# Population immunity to clade 2.3.4.4b H5N1 is dominated by anti-neuraminidase antibodies

**DOI:** 10.64898/2026.02.10.26346014

**Authors:** Gagandeep Singh, Disha Bhavsar, Enikö Hermann, Charles Gleason, Harkirat Singh Sandhu, Parul Singh, Jessica Nardulli, Neko Lyttle, Yuexing Chen, Suzana Sabaiduc, Garazi Peña Alzua, Danuta M Skowronski, Viviana Simon, Florian Krammer

**Author notes:** Corresponding Author: Florian Krammer.

## Abstract

Clade 2.3.4.4b highly pathogenic avian influenza A(H5N1) viruses continue to expand geographically and across mammalian hosts, raising concern about pandemic potential. The degree and specificity of pre-existing immunity in humans are key determinants of this risk. We analyzed hemagglutinin (HA)-and neuraminidase (NA)-specific antibody responses in 300 sera collected from adults in New York City. While HA directed binding antibodies to clade 2.3.4.4b H5 were low and hemagglutination-inhibiting antibodies were absent, we detected widespread binding and functional NA antibodies against N1 neuraminidases from clade 2.3.4.4b H5N1 viruses. Neuraminidase inhibition (NI) titers were highest against North American D1.1 genotype N1 viruses and correlated strongly with neutralizing activity, whereas HA-binding antibodies did not. An additional N-linked glycosylation site, as found in the NA of a human D1.1 isolate from British Columbia, reduced susceptibility to NI antibodies. Antibodies to N5 from H5N5 were minimal. These findings indicate that population-level immunity to clade 2.3.4.4b H5 viruses is dominated by NA-directed antibodies, with important implications for pandemic risk assessment.

**Importance:** Understanding how pre-existing human immunity shapes susceptibility to emerging influenza viruses is central to pandemic preparedness. Here, we determined that human sera contain widespread, functional antibodies targeting H5N1 neuraminidase, which correlate with virus neutralization, whereas HA directed responses are limited. We further show that acquisition of an NA glycosylation site reduces antibody inhibition, highlighting a potential pathway for immune evasion. These results identify neuraminidase-specific immunity as a major immunological barrier to severe H5N1 disease in humans and emphasize the need to incorporate NA antigenicity into influenza surveillance, risk assessment, and next-generation vaccine design.

## Introduction

H5N1 highly pathogenic avian influenza (HPAI) viruses have been described as early as 1959(1) with the first documented human cases, including some with fatal outcomes, being described starting 1997(2, 3). More recently, a subclade of the A/goose/Guangdong/1/1996(4) lineage of H5N1, clade 2.3.4.4b H5N1 has caused a panzootic and is now present in all continents, with the exception of Oceania, in many wild avian populations(5, 6). It also causes frequent infections of marine mammals(7–9), mammalian terrestrial predators, and scavengers(10). The virus has also caused dramatic losses in the poultry industry, has started to spread in US dairy cattle(11, 12), and has infected mice, cats and dogs. Finally, approximately 70 infections of humans with this new H5N1 clade have been detected, most of them mild but severe cases and fatalities have been reported as well(5, 13). Importantly, clade 2.3.4.4b H5N1 has reassorted with different North American avian influenza viruses(14–16). While these reassortant viruses all carry the clade 2.3.4.4b H5 hemagglutinin (HA), they may express Eurasian avian N1, North American avian N1 (D1.1 genotype)(17), as well as N5(18) or N8(19) neuraminidase (NA). Of note, most severe or fatal human cases have either been caused by the D1.1 genotype(20) or, in one instance, by H5N5(21).

The human population is constantly exposed to seasonal H1N1, H3N2 and type B influenza viruses through infections and vaccination. Individuals born before 1968 may also have been exposed to the H2N2 subtype which circulated between 1957 and 1968(22). This exposure has induced pre-existing immunity to influenza A viruses in humans. T-cell responses to conserved epitopes of internal proteins like the nucleoprotein or the matrix protein are common and can likely recognize H5 infected cells as well(23, 24). In addition, while phylogenetically distinct, the N1 NA found in current seasonal human H1N1 strains is from the same NA subtype as the N1 NAs found in H5N1. Finally, both H1 and H5 HA are group 1 HAs and it is likely that antibodies to conserved sites on HA, for example, the stalk domain, can cross-react to H5 HA as well. Here we set out to measure these cross-reactive HA and NA responses using a panel of 300 human sera collected between February 2024 and April 2025 representing the general adult population of a large metropolitan city in North America (New York City) ranging in age from 18 to 89 years and born between 1940 and 2006 (**Table S1**).

## Results

### Binding antibody responses to clade 2.3.4.4b H5 HA

First, we measured binding antibodies to clade 2.3.4.4b HA for all 300 human sera using an enzyme-linked immunosorbent assay (ELISA). The H5 strain chosen for this assay was A/California/135/2024 (B3.13 genotype) isolated from a human infection. As comparators, we used H1 HA from the 2009 pandemic strain A/California/04/2009 to which all subjects should have antibodies. We also included H5 HA from an older H5 strain belonging to clade 1 A/Vietnam/1203/2004 as well as from an H7 strain (A/Guangdong/17SF003/2016). The H7 HA was included since we have historically seen that cross-reactivity to H7 HA is one of the lowest in humans among all HA subtypes(25). Comparative sequence analysis demonstrated substantial divergence between the HA of clade 2.3.4.4b A/California/135/2024 (H5N1) and A/California/04/2009 (H1N1) viruses, with approximately 63% amino-acid identity and multiple differences across defined antigenic sites (**Table S2**). As expected, reactivity was highest (**Figure 1A**) to the H1 HA (GMT 2797.2), followed by reactivity to the older H5 HA (GMT, 787.1, p< 0.0001 vs H1N1). Reactivity to clade 2.3.4.4b H5 HA was lower (GMT 32.5, p< 0.0001 vs H1N1) but remained higher than that observed for H7 HA (GMT 67, p< 0.0001 vs H1N1). When we stratified titers by age, we could observe that H1 reactivity was highest in younger people (**Figure 1B**). This is expected since younger individuals would have been exposed to this specific H1 strain early in life leading to strong immune responses or antigenic imprinting. For the historic H5 HA and clade 2.3.4.4b HA reactivity was stronger in individuals born before 1968 (**Figure 1C and D**) who may have higher stalk-reactive antibodies to group 1 viruses due to imprinting to H2N2 and H1N1 viruses. Finally, H7-specific antibody titers were generally low but were highest among individuals born in the 1960s, potentially reflecting effects of H3N2 imprinting (**Figure 1E**).

**Fig. 1.**
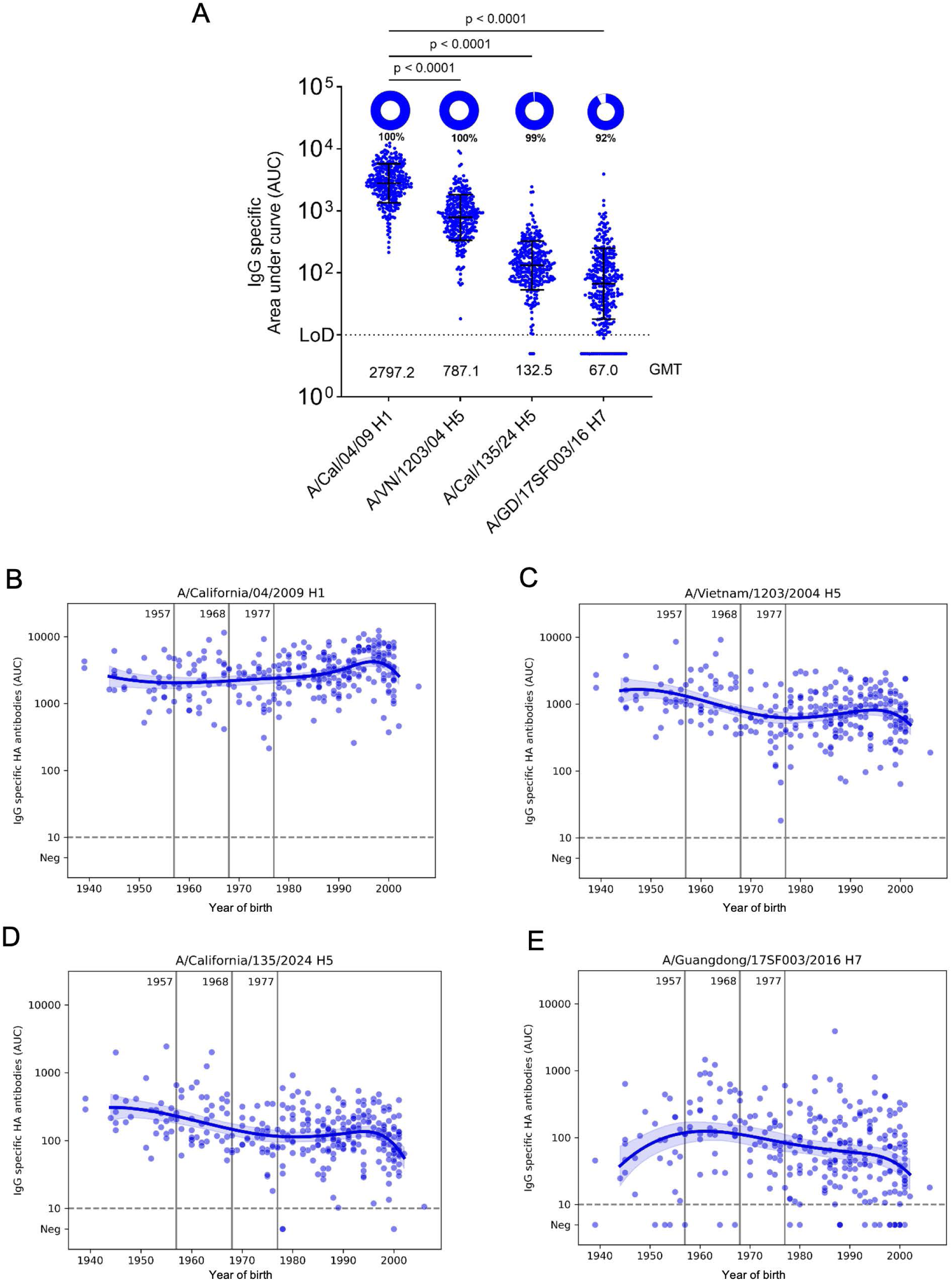
Population-level HA-binding antibody responses in 300 human sera from New York City. **(A)** Immunoglobulin G (IgG) binding antibodies were quantified by enzyme-linked immunosorbent assay (ELISA) against hemagglutinin (HA) recombinant proteins from influenza A/California/04/2009 (H1N1), A/Vietnam/1203/2004 (H5N1), clade 2.3.4.4b A/California/135/2024 (H5N1), and A/Guangdong/17SF003/2016 (H7N9). Titers are shown as area under the curve (AUC) with the geometric mean and geometric standard deviation denoted by error bars. Limits of detection (LOD) are shown as dotted lines on each graph. The percentage of serum samples for each antigen with AUC values above the LOD are shown as pie charts, with the percentage of positive samples displayed below the pie chart. GMT, geometric mean titer for each antigen were displayed in graph. Binding titers across different HAs were compared using Kruskal–Wallis test and Dunn’s multiple comparison test; *P* values are indicated above brackets. **(B-E),** HA-binding antibody titers stratified by year of birth. Vertical lines denote the emergence of the 1957 H2N2, 1968 H3N2, and 1977 H1N1 influenza viruses. Each circle represents the geometric mean binding antibody titer from an individual serum sample; locally estimated scatterplot smoothing curves are shown.

### Binding antibody responses to avian N1 and N5 NAs associated with clade 2.3.4.4b HA

Next, we assessed binding antibody responses to relevant NAs that are associated with clade 2.3.4.4b H5 by ELISA. Here, we included Eurasian avian N1 from H5N1 strains A/bald eagle/FL/W22-134-OP/2022 (B1.1 genotype) and A/California/135/2024 (B3.13 genotype) and North American avian N1 from a genotype D1.1 isolate from a fatal human case, A/Louisiana/12/2024 (13). We also included an N5 antigens associated with clade 2.3.4.4b H5N5 (A6 genotype) from A/great black-backed gull/NS/FAV-0405-1/2023 and A/Washington/2148/2025, the latter virus associated with 2025 fatal human case in Washington State (26). As a positive control, the N1 of human A/California/04/2009 H1N1 was included. Sequence analysis revealed higher amino-acid identity (78-84%) between the N1 neuraminidases of clade 2.3.4.4b A(H5N1) viruses and recently circulating seasonal A/California/04/2009 (H1N1) virus (**Table S3**). We also measured antibodies to the N1 NA of an older H5N1 strain, A/Vietnam/1203/2004 and from an N9 NA from the A/Anhui/1/2013 H7N9 strain. N1 reactivity was lowest to A/Vietnam/1203/2004 N1 (**Figure 2A**; GMT 78.9). Interestingly, the reactivity to N1 from human H1N1 (GMT 181.6) and reactivity to the N1 NAs from strains A/bald eagle/FL/W22-134-OP/2022 (GMT 103.4, p = 0.0011 vs H1N1) and A/California/135/2024 (GMT 123.4, p = 0.1229 vs H1N1) were very similar. Reactivity to the N1 NA from the D1.1 genotype (A/Louisiana/12/2024) was higher (GMT 306.8, p= 0.0220 vs H1N1) than that for the human H1N1 NA. In contrast, reactivity to N5 A/great black-backed gull/NS/FAV-0405-1/2023 (GMT 5, P< 0.0001 vs H1N1) and A/Washington/2148/2025 (GMT 21.4, P< 0.0001 vs H1N1) as well as to the N9 antigen (GMT 20.6) was low or absent in most individuals. When stratified by birth year, reactivity to N1 from human H1N1 was highest in the population born around 2000 as expected (**Figure 2B**). Titers to N1 from the older H5N1 strain were slightly higher in the older and younger individuals, with a noticeable shift occurring around birth years in the late 1980s. (**Figure 2C**). Binding titers to the N1 NAs of A/bald eagle/FL/W22-134-OP/2022 and A/California/135/2024 appeared to be very similar and showed only a weak suggestion of a W-shaped pattern, with the highest titers primarily observed at the youngest and oldest age extremes, with a modest increase among those born in the early 1970ies (**Figure 2D and E**). Binding to the N1 of the D1.1 genotype virus A/Louisiana/12/2024 was also highest in the oldest and youngest study participants but lacked the increased binding in the 1970ies birth cohort (**Figure 2F**). In contrast, very low or negligible levels of binding antibodies were detected across individuals from different birth years against N5 A/great black-backed gull/NS/FAV-0405-1/2023 and A/Washington/2148/2025 (**Figure 2G and 2H**). Finally, binding to N9 was low but a peak in binding was recognizable in individuals born in the 1980ies and 90ies (**Figure 2I**).

**Fig. 2.**
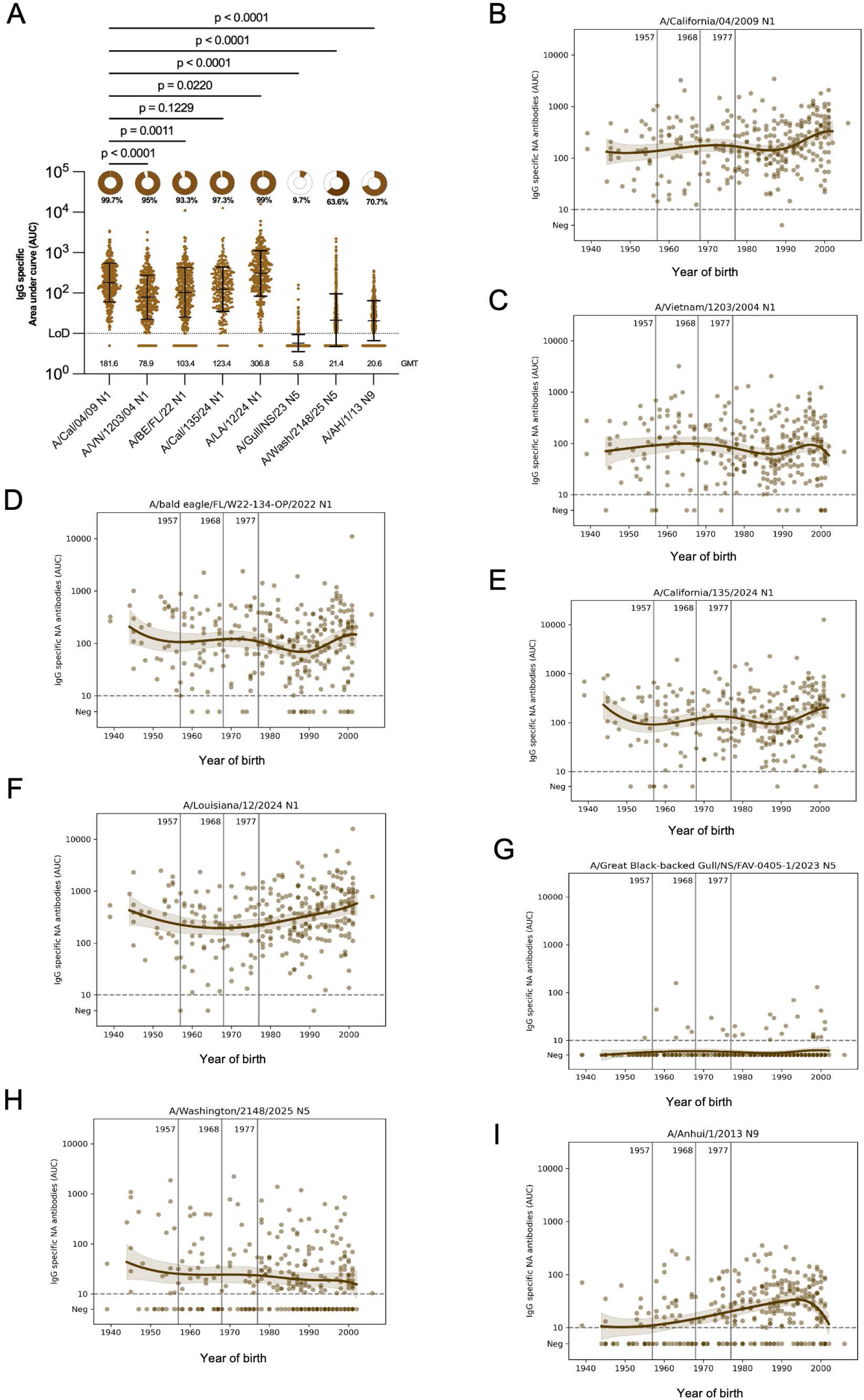
Population-level NA-binding antibody responses in 300 human sera from New York City. (**A)** Immunoglobulin G (IgG) binding antibodies were quantified by enzyme-linked immunosorbent assay (ELISA) against neuraminidase (NA) recombinant proteins from influenza A/California/04/2009 (H1N1), A/Vietnam/1203/2004 (H5N1), clade 2.3.4.4b A/bald eagle/FL/W22-134-OP/2022 (H5N1), A/California/135/2024 (H5N1, B3.13 genotype), A/Louisiana/12/2024 (H5N1, D1.1 genotype), A/great black-backed gull/NS/FAV-O405-1/2023 (H5N5) and A/Anhui/1/2013 (H7N9). Titers are shown as area under the curve (AUC) with the geometric mean and geometric standard deviation denoted by error bars. Limits of detection (LOD) are shown as dotted lines on each graph. The percentage of serum samples for each antigen with AUC values above the LOD are shown as pie charts, with the percentage of positive samples displayed below the pie chart. GMT, geometric mean titer for each antigen were displayed in graph. Binding titers across different NAs were compared using Kruskal–Wallis test and Dunn’s multiple comparison test; *P* values are indicated above brackets. **(B-H),** NA-binding antibody titers stratified by year of birth. Vertical lines denote the emergence of the 1957 H2N2, 1968 H3N2, and 1977 H1N1 influenza viruses. Each circle represents the geometric mean binding antibody titer from an individual serum sample; locally estimated scatterplot smoothing curves are shown.

### Hemagglutination inhibition (HI) and microneutralization titers to clade 2.3.4.4b H5N1

After assessing binding antibody levels, we also wanted to determine functional antibodies to clade 2.3.4.4b H5N1. Again, we used the A/California/04/2009 H1N1 strain as control. First, we ran HI assays. The HI titer is a well-established correlate of protection for seasonal influenza viruses(27, 28) and it measures antibodies to the head domain of the HA that can block receptor binding. As expected, the majority of individuals had HI titers to the H1N1 strain with a geometric mean titer (GMT) of 18.7 but HI titers to clade 2.3.4.4b A/bald eagle/FL/W22-134-OP/2022 H5N1 (B1.1 genotype) were undetectable (**Figure 3A**). We also ran a microneutralization assay with both viruses. This assay is a multicycle assay with serum in the liquid overlay at all times(29) and can therefore also measure the impact of anti-NA and anti-HA stalk antibodies that cannot be detected in HI assays. Here, titers to H1N1 (GMT 30. 5) were higher but activity to clade 2.3.4.4b H5N1 (GMT 15.4) was at or above the limit of detection for most individuals (**Figure 3B**). We then also stratified the HI and microneutralization data by birth year. HI titers to H1N1 were highest in individuals born around the year 2000 (**Figure 3C**). Since no HI titers were detected to H5N1, the age stratification just showed a flat line (**Figure 3D**). For the microneutralization assay, titers to H1N1 followed the same trend as the HI titers with the youngest individuals having higher titers (**Figure 3E**). A similar trend, although at a lower level, was seen when the H5N1 microneutralization titers were age stratified (**Figure 3F**), suggesting the role of anti-HA stalk or anti-NA antibodies in inhibiting viral replication *in vitro*.

**Fig. 3.**
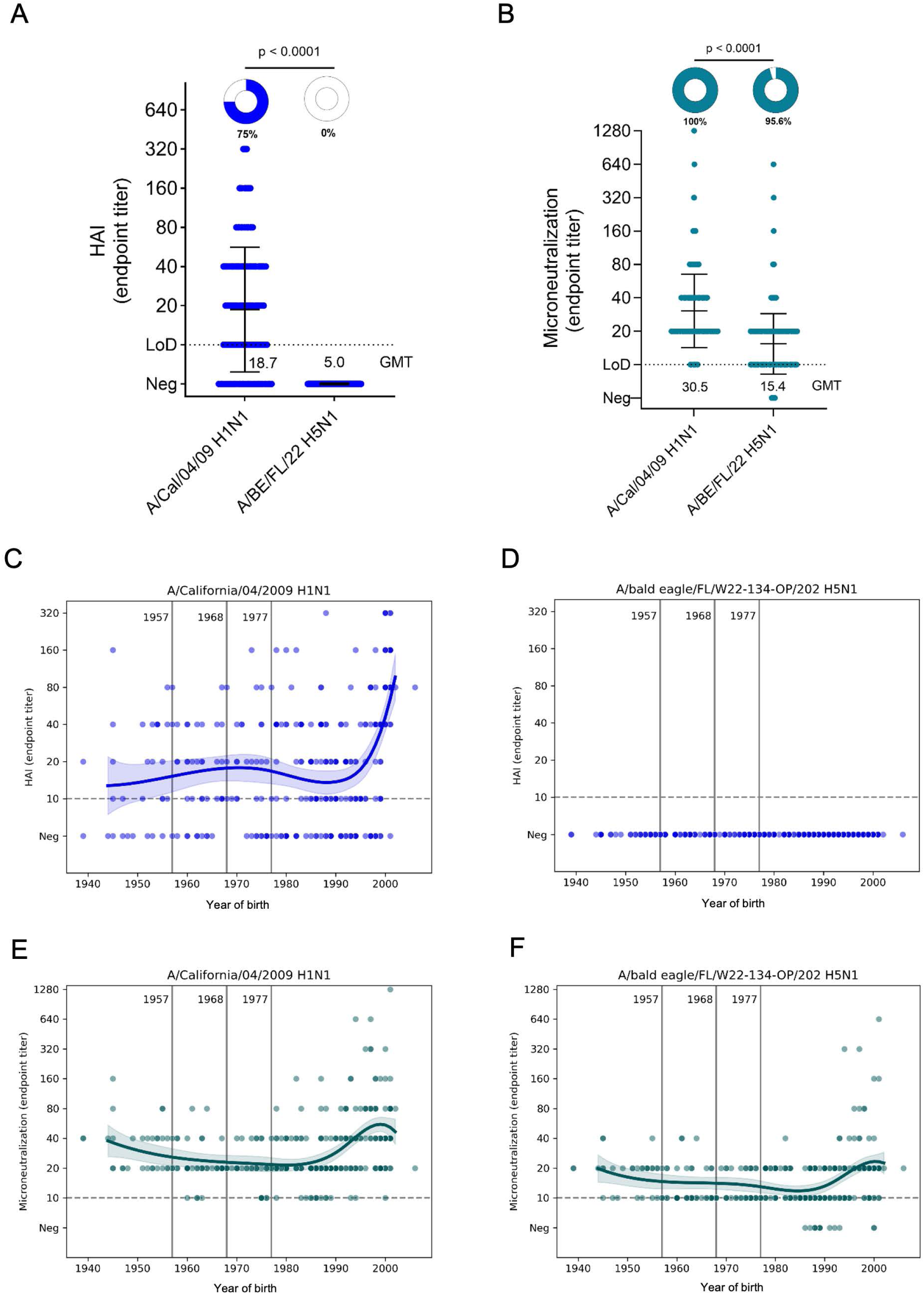
Hemagglutination inhibiting (HI) and neutralizing antibody responses against H1N1 and clade 2.3.4.4b H5N1 viruses in 300 human sera from New York City. (**A**) HI antibody response of human sera against influenza A/California/04/2009 (H1N1) and clade 2.3.4.4b A/bald eagle/FL/W22-134-OP/2022 (H5N1). HAI titers were presented as HAI endpoint titers and geometric mean titers (GMT) with error bars depicting standard deviation (SD). The assay detection limit for both the assays were 1:10, and samples with antibody below the detection limit were assigned to an arbitrary antibody titer of 5, which is used to calculate geometric mean titer. Limits of detection (LOD) are shown as dotted lines on each graph. The percentage of serum samples for each virus with end titer above the LOD are shown as pie charts, with the percentage of positive samples displayed below the pie chart. GMT, geometric mean titer for each antigen were displayed in graph. (**B**) Multicycle neutralization assay was performed to measure antibody responses against influenza A/California/04/2009 (H1N1) and clade 2.3.4.4b A/Bald eagle/FL/W22-134-OP/2022 (H5N1). Limits of detection (LOD) are shown as dotted lines on each graph. The percentage of serum samples for each virus with end titer above the LOD are shown as pie charts, with the percentage of positive samples displayed below the pie chart. GMT, geometric mean titer for each antigen were displayed in graph. Statistical significance of differences between the two different viruses were calculated using a Kruskal-Wallis test followed by a Dunn’s multiple-comparisons test; *P* values are indicated above brackets. (**C-F**) HI and microneutralization antibody titers stratified by year of birth. Vertical lines denote the emergence of the 1957 H2N2, 1968 H3N2, and 1977 H1N1 influenza viruses. Each circle represents the geometric mean binding antibody titer from an individual serum sample; locally estimated scatterplot smoothing curves are shown.

### Neuraminidase inhibition titers to clade 2.3.4.4b H5N1 viruses

After assessing functional HA and neutralization titers, we also wanted to investigate functional NA titers using a neuraminidase inhibition (NI) assay. NI antibodies are also an established correlate of protection for seasonal influenza viruses (30–32). In order to measure NA-based NI without interference from anti-HA antibodies, viruses with HAs to which the human population is not exposed to are usually employed. For A/bald eagle/FL/W22-134-OP/2022 (B1.1 genotype) and A/Vietnam/1203/2004 we had already H5N1 viruses but for A/California/04/2009, A/Louisiana/12/2024 (D1.1 genotype) and A/California/135/2024 (B3.13 genotype) we generated mismatched H15N1 viruses. Of note, H5 versus H15 HA could still influence NI activity and therefore, comparisons between the H15Nx strains are most valuable. Titers were lowest to the A/Vietnam/1203/2004 strain (GMT 15.1, H5), followed by A/California/135/2024 (GMT 27.8, H15) (**Figure 4A**). Titers to A/bald eagle/FL/W22-134-OP/2022 (GMT 68.5, H5) and A/California/04/2009 (GMT 38.2, H15) were higher and of comparable magnitude. The highest titer was measured against A/Louisiana/12/2024 (GMT 80.0, H15) which tracks well with antibody binding titers in ELISA which were also highest against this strain. Titers to N5 from A/great black-backed gull/NS/FAV-0405-1/2023 (GMT 13.0, H15) and A/Washington/2148/2025 (GMT 14.6, H15) were minimal in most of the individuals. The slightly higher binding for the A/Washington/2148/2025 N5 may be due to the fact that is has one N-linked glycosylation site less than the N5 from A/great black-backed gull/NS/FAV-0405-1/2023. When stratified by birth year, titers in general followed the same pattern across viruses. Titers were highest among the oldest and youngest individuals (**Figure 4B to F**).

**Fig. 4.**
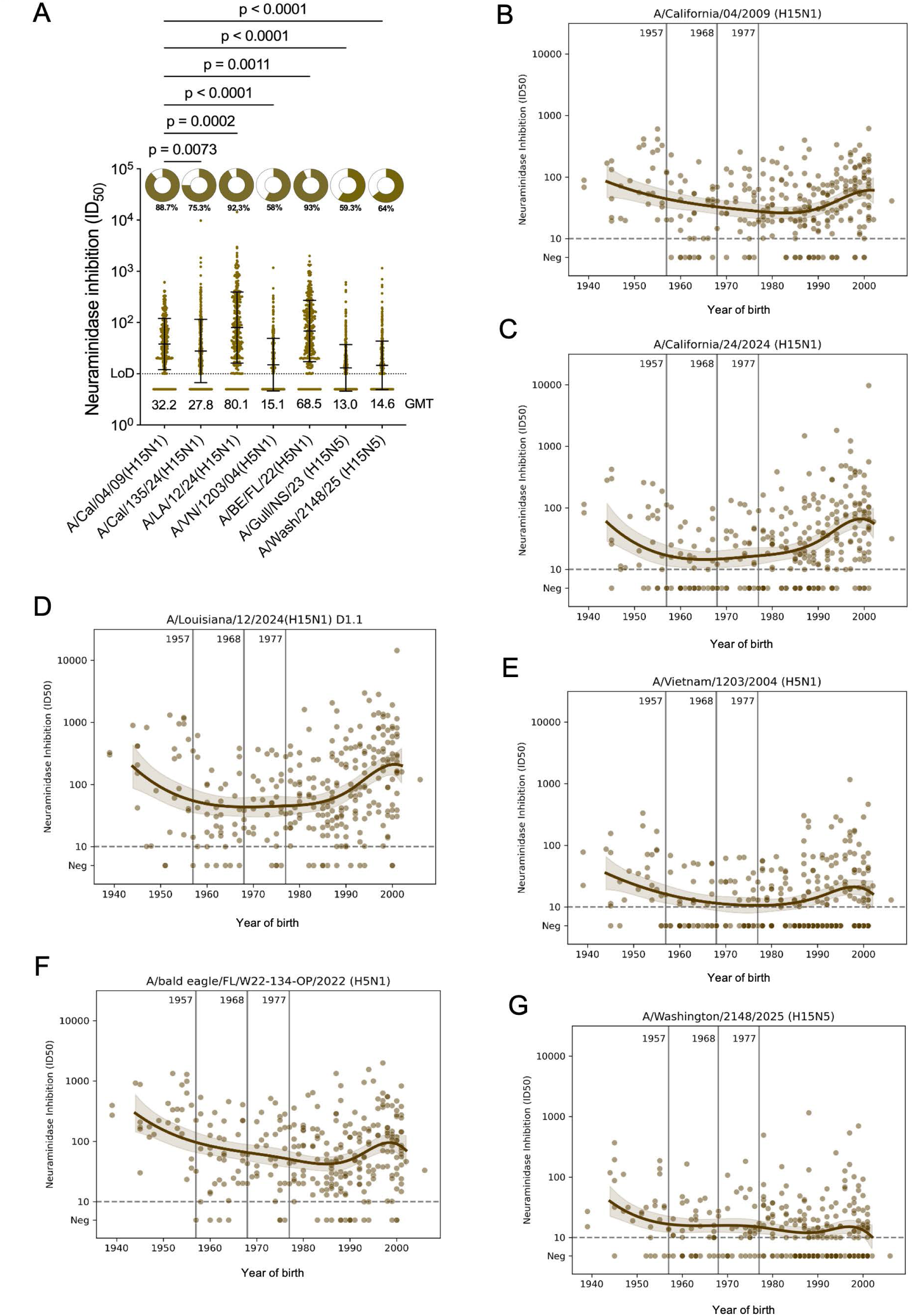
Functional neuraminidase inhibition (NI) antibody responses to N1 of H1N1 and clade 2.3.4.4b H5N1 viruses in 300 human sera from New York City. **(A)** NI antibody responses were measured by enzyme linked lectin assay (ELLA) using A/bald eagle/FL/W22-134-OP/2022 (H5N1), A/Vietnam/1203/2004 (H5N1) and H15N1 viruses with mismatch HA and target NAs from A/California/04/2009, A/Louisiana/12/2024 and A/California/135/2024. Limits of detection (LOD) are shown as dotted lines on each graph. The percentage of serum samples for each virus with IC_50_ values above the LOD are shown as pie charts, with the percentage of positive samples displayed below the pie chart. GMT, geometric mean titer for each antigen were displayed in graph NI titers across different N1 were compared using Kruskal–Wallis test and Dunn’s multiple comparison test; *P* values are indicated above brackets. (**B-F**) NI titers stratified by year of birth. Vertical lines denote the emergence of the 1957 H2N2, 1968 H3N2, and 1977 H1N1 influenza viruses. Each circle represents the geometric mean binding antibody titer from an individual serum sample; locally estimated scatterplot smoothing curves are shown.

### NA specific antibodies correlate with multicycle microneutralization titers

Given the low probability of prior H5N1 exposure in general population, the presence of neutralizing antibodies was unexpected and suggested contributions from antibody targets beyond the HA head, including NA and the conserved HA stalk domain. To assess the relative contributions of these targets, we performed correlation analyses between the A/bald eagle/FL/W22-134-OP/2022 microneutralization titers and different NA and HA-specific antibody responses. We observed a strong correlation between H5N1 neutralizing antibodies and functional NI antibodies of clade 2.3.4.4b N1 viruses (Spearman’s *r* ranging from 0.50-0.88) (**Figure S1A-D**). Likewise, we noted a strong positive correlation between NA specific binding antibodies levels and H5N1 neutralization titers (**Figure S1E-H**) in general. In contrast, no significant correlation was detected between HA-specific IgG binding levels and H5N1 neutralization (**Figure S1I**).

### Effect of an additional glycosylation site in the N1 NA of a human D1.1 isolate

In 2024, a clade 2.3.4.4b D1.1 H5N1 strain caused a severe human infection in British Columbia(20). Sequences from the patient isolate suggest, that the virus acquired an additional putative N-linked glycosylation site in its N1 NA in position 270 (N1 numbering) (**Figure 5A**). Phylogenetic analysis of the NA of the sequence suggests, that the viruses most closely related to this isolate did already have this glycosylation site (**Figure 5B and C**). Of course, this finding triggered the question if this putative glycosylation site impacts on human NA immunity. To assess this, we ran a side-by-side NI assay with an H15N1 reassortant carrying the NA of A/Louisiana/12/2024 (D1.1 N1, lacking the glycosylation site) and an H15N1 reassortant carrying the A/British Columbia/PHL-2032/2024 NA (D1.1 N1, with the additional glycosylation site). Our analysis showed that NI titers were substantially lower against the virus with the additional glycosylation site compared to the virus without it, with GMTs of 57.2 for A/British Columbia/PHL-2032/2024 versus 98.0 for A/Louisiana/12/2024, respectively. (**Figure 5D**).

**Fig. 5.**
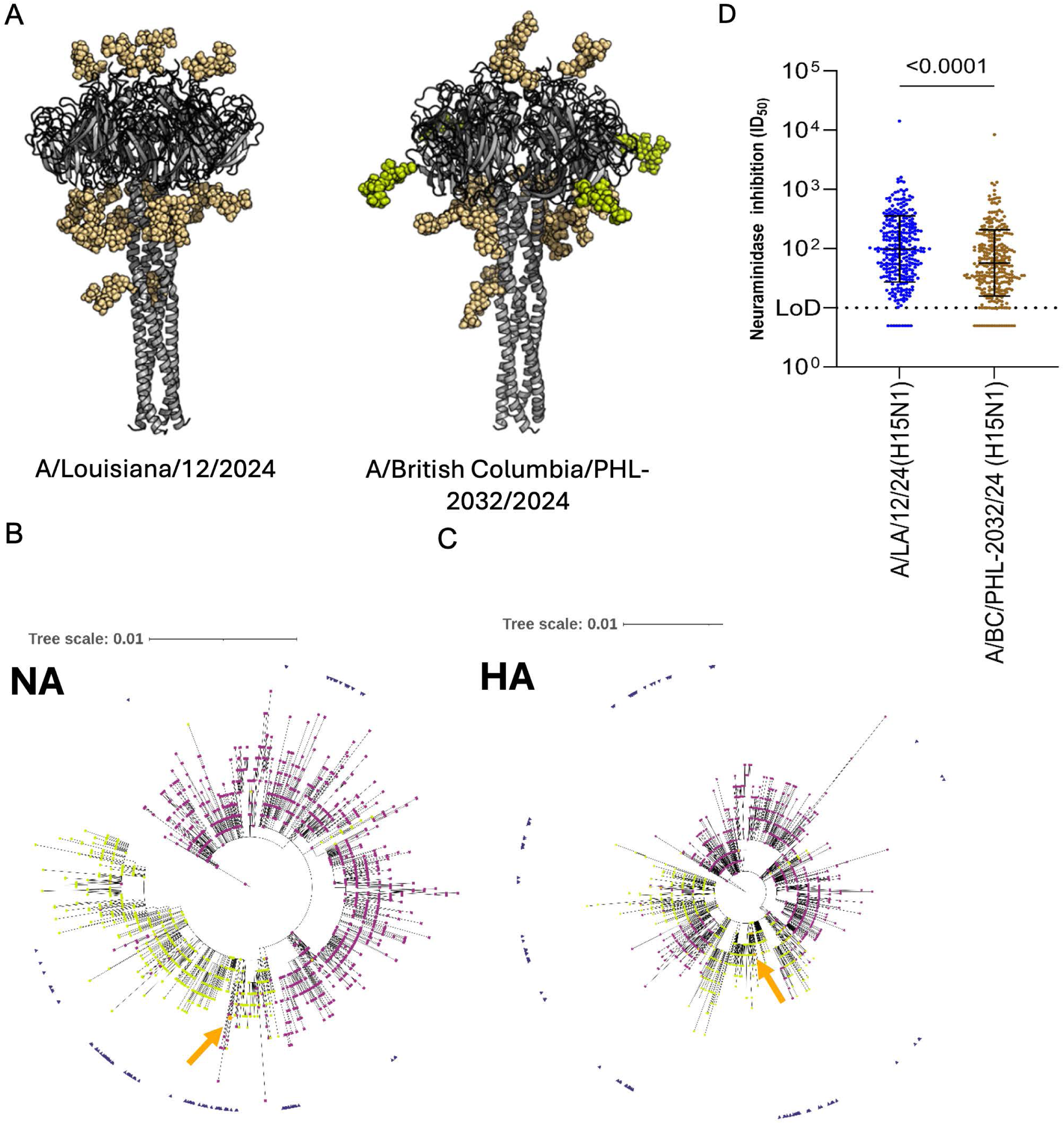
Additional N-linked glycosylation in the N1 neuraminidase (NA) of a human D1.1 isolate. **(A)** Structural models of NA from two human D1.1 H5N1 isolates showing predicted N-linked glycosylation sites (spheres). The additional glycosylation site in A/British Columbia/PHL-2032/2024 is highlighted in lime green. (**B and C**) Phylogenetic analysis of H5N1 clade 2.3.4.4b genotype D1.1 HA and NA nucleotide sequences available in GISAID. Trees were generated using FastTree (NGPhylogeny.fr) and visualized with iTOL. Viruses with a potential N-linked glycosylation site at NA residue 270 (N1 numbering) are indicated by lime green circles; those lacking the site are shown as purple squares. The human isolate A/British Columbia/PHL-2032/2024 is highlighted in orange and indicated with an orange arrow, and samples from the same Pacific Northwest region from the October-December 2024 timeframe are marked with blue triangles on the outside. (**D**) NI antibody responses measured by ELLA using H15N1 viruses bearing mismatched HA and NAs from A/Louisiana/12/2024 or A/British Columbia/PHL-2032/2024. Statistical significance was assessed using the Kruskal Wallis test with Dunn’s multiple-comparison correction; *P* values are show

## Discussion

How much pre-existing immunity exists in humans to animal influenza virus strains with pandemic potential is a very important question. High levels of cross-reactive antibodies or T-cells are a potential barrier to efficient virus spread in the human population. In the best-case scenario, cross-reactive immunity may prevent a pandemic. But even if these cross-reactive responses are not powerful enough to prevent a pandemic, they could still significantly ameliorate disease severity and overall impact of a new pandemic as seen in 1968 with NA antibodies(33) and in 2009 with HA and NA antibodies. Several studies have already investigated cross-reactive immune responses to clade 2.3.4.4b H5N1 (24, 34–38) and have, most prominently, identified high N1 cross-reactive antibody levels (14, 24, 39–41). Our data align well with published reports in general, suggesting high N1 cross-reactive and functional responses but lower HA-based immunity with limited functionality. However, there are several aspects of our data that stand out and/or significantly add new information.

The reactivity patterns that we observe in our assays broadly align with studies by Skowronski *et al.*(41), Ort *et al.*(40), Garretson *et al.* (36), and Li *et al.* (37). Similar to Garretson *et al.* we see the highest H5 HA binding antibodies in older individuals who may have been exposed to H1 and H2 HAs and may therefore have higher stalk-reactive antibody titers (36). In fact, we have previously shown much higher anti-HA stalk antibody titers and antibody titers to clade 2.3.4.4 HA in older individuals than in middle-aged or younger individuals (42). Of note, overall, we saw much lower binding titers to clade 2.3.4.4b H5 HAs than for a historic H5 HA. This could be due to real differences in binding or due to stability issues with clade 2.3.4.4b HAs which in our hands tend to aggregate.

NA reactivity by birth year also aligns well with published data. While we did not have very young individuals in our study, we certainly see both the peak in individuals born in 1990-2000 as well as the peak in titers in very old individuals for binding NA and NI titers that Skowronski *et al.*, Ort *et al.* and *Li et al.* describe as well.

Our results for HI antibodies align well with published studies – suggesting an absence of any HI active antibodies to clade 2.3.4.4b H5 in the human population. However, we did detect low but significant neutralization activity against A/bald eagle/FL/W22-134-OP/2022 (H5N1, B1.1 genotype). This differs from results by Li *et al.* who saw no neutralizing activity but aligns well with other reports that showed low but detectable neutralizing activity. The differences in neutralization activity may arise from different assays used. Zhang *et al.*(34) and Daniel *et al.*(35) used pseudotyped virus particle entry inhibition assays and saw low neutralization activity. The pseudotyped virus particle entry inhibition assay is much more sensitive to stalk-reactive antibodies ((43) as well as supplementary figure 2 in(44)) and often overreports which explains these results. The neutralization assay used by Li *et al.*(37) which did not detect any neutralization is based on authentic virus entry inhibition without a multicycle stage which is known for being relatively insensitive for stalk-based neutralization and NA-based inhibition. We and Garretson *et al.*(36) used multicycle assays which are typically more sensitive to anti-stalk antibodies but also detect NA-based immunity. Given the high titers of NI active NA antibodies, it is very likely that the majority of the neutralization activity seen by us is mediated by anti-NA antibodies.

What makes our study unique, is the focus on D1.1 genotype in terms of NA immunity. We did see higher NI titers to D1.1, which carries a North American avian N1 compared to other tested 2.3.4.4b clade H5N1 isolates that carry a Eurasian avian N1 when the same mismatched H15 HA was used. Furthermore, we investigated the impact of an additional putative N-linked glycosylation site in the N1 NA that some D1.1 isolates carry, including the one causing the severe case in British Columbia(20). This additional putative glycosylation site did increase resistance to anti-NA antibodies in human sera. Viruses carrying that glycosylation site may be less susceptible to pre-existing N1-directed immunity and might therefore pose higher infection and/or severity risk or may lead to longer shedding.

Interestingly, we also found lower NI titers to the older clade 1 A/Vietnam/1203/2004 as compared to the clade 2.3.4.4b viruses. Older H5N1 viruses, including the Vietnam strain, have a deletion in the stalk domain of the NA(45). It has been proposed that the deletion in the NA stalk is a pathogenicity factor in poultry but that a longer stalk, as present in clade 2.3.4.4b H5N1 (including in the D1.1 genotype) is a prerequisite for transmission in mammals. However, it could be that the longer stalk makes the NA more vulnerable to NI active antibodies present in humans.

Historically, H5N1 infections in humans have been associated with very high case fatality rates of approximately 50%(46) (even though infection fatality rates have been reported to be much lower(47)). This does not align with case fatality rates observed with clade 2.3.4.4b H5N1 viruses where most detected infections were mild, even though a small number of severe cases and deaths have been reported as well(46). While the reason for this may be multifactorial, it stands to reason that the high NA binding antibody titers and NI titers to these viruses may be one of the reasons for the apparent lower pathogenicity in humans (in contrast to naïve mammals like cats, foxes, bears, seals and ferrets, where clade 2.3.4.4b viruses are very pathogenic). In fact, ferrets pre-infected with H1N1 virus showed high resistance to clade 2.3.4.4b H5N1 challenge in comparison to H3N2 or influenza B virus pre-infected animals(48, 49). Only H1N1 pre-infected ferrets had cross-reactive anti-NA antibodies to H5N1(48, 49). This aligns with the fact that NA antibodies represent a correlate of protection in humans(30–32). Historically, pre-existing anti-N2 antibodies have also been shown to have provided protection against H3N2 during the pandemic of 1968(33). If the N1 antibody titers in the population would be high enough to be a barrier for H5N1 to become a pandemic is unclear. But it stands to reason that the substantial NA-Cross-reactivity to both the Eurasian avian N1 (present in most clade 2.3.4.4b H5N1 strains) and to the North American NA (present in D1.1 H5N1 strains) could substantially ameliorate severity of an H5N1 pandemic. Recently, clade 2.3.4.4b H5N5 viruses emerged in Canada and also caused a fatal human infection in Washington State. In our study we also assessed immunity to the N5 from these strains and found very little cross-reactivity. It seems therefore that H5N5 would have less of an immunological barrier to become a pandemic than H5N1 and its impact may also be more severe.

Our study has several limitations. First, the cohort is skewed toward younger adult and female participants, which may modestly affect estimated antibody levels relative to the general population. Second, although we assessed multiple antibody targets, other aspects of adaptive immunity, including cell-mediated immune responses that can contribute to reduced influenza disease severity, were not examined here. Third, only a small number of strains was included which may limit generalizability of the conclusions.

## Materials and Methods

### Experiment model and study participants details

Sera were collected between February 2024 and April 2025 from adult study participants who live or work in New York City as part of the observational study APOLLO (Antibody panels of longitudinal levels of coronavirus immunity) carried out at the ISMMS (Icahn School of Medicine at Mount Sinai). The study is approved by the Mount Sinai Hospital Institutional Review Board (STUDY-23-00873) and recorded as study protocol DMID 24-0011 as part of the Center for Research on Influenza Pathogenesis (CRIPT). All participants signed written consent forms prior to sample and data collection. We selected sera from adult study participants between 18 to 89 years of age at the time of sera collection (214 female, 84 male, 2 missing sex data, total: 300 unique participants) (**Figure S2 and Table S1**). Vaccination history was not a criteria for sample selection.

### Cell lines, Viruses and recombinant proteins for immunological assays

Sf9 cells (CRL-1711, ATCC) used for baculovirus rescue were cultured in *Trichoplusia ni* medium-formulation Hink insect cell medium (TNM-FH, Gemini Bioproducts), supplemented with 10% fetal bovine serum (FBS, Gibco) and antibiotics (100 units/mL penicillin–100 µg/mL streptomycin [Pen-Strep]; Gibco) and 0.1% Pluronic F-68 (Gibco). High Five cells (BTI-TN-5B1-4, B85502, Thermo Fisher Scientific), utilized for recombinant HA and NA production, were grown in serum-free Sf-900 medium (Gibco) supplemented with Pen-Strep. Madin-Darby canine kidney (MDCK) cells were grown in Dulbecco’s Modified Eagle Medium (DMEM) (Gibco) containing Pen-Strep antibiotics mix and 10% FBS resulting in complete DMEM (cDMEM). MDCK cells were maintained at 37 °C with 5% CO_2_.

Influenza A viruses were grown in 8–10 day old embryonated chicken eggs (Charles River Laboratories) at 37 °C for 48 hours (h) and cooled at 4°C overnight (O/N). Virus reassortants were rescued by plasmid-based reverse genetic techniques as previously described(50). A/bald eagle/FL/W22-134-OP/2022 (H5N1, 6:2 A/PR/8/1934) and A/Vietnam/1203/2004 (H5N1, 6:2 A/PR/8/1934) viruses were rescued with the HA and NA from the original strain and the remaining six segments from A/PR/8/34 H1N1 via reverse genetics, with removal of the H5 polybasic cleavage site. H15N1_A/California/04/2009,_ H15N1_A/Louisiana/12/2024,_ H15N1_A/California/135/2024_ H15N1_A/British Columbia/PHL-2032/2024,_ H15N5 _A/great black-backed gull/NS/FAV-0405-1/2023 /12/2024_ and H15N5 _A/Washington/2148/2025_ were rescued with hemagglutinin (HA) H15 from the A/wt shearwater/WA/2576/1979 (H15N9) and neuraminidase (NA) N1 from A/California/04/2009 (H1N1), A/Louisiana/12/2024 (H5N1), A/California/135/2024 (H5N1), A/British Columbia/PHL-2032/2024 (H5N1), N5 from A/great black-backed gull/NS/FAV-0405-1/2023 (H5N5) and A/Washington/2148/2025 (H5N5) respectively and the remaining six segments from A/PR/8/34 as 6:2 reassortant virus. The A/PR/8/34 backbone is attenuated in humans, ferrets and chickens, does not support efficient transmission in guinea pigs and is commonly used as a safe laboratory backbone for reassortant influenza viruses(51–55). Assays using H15NX reverse genetics viruses were conducted in biosafety level 2+ laboratories.

All recombinant HA and NA proteins were expressed using the baculovirus expression system as described in detail previously(56–58). Recombinant HA proteins (as soluble trimers) used in this study were derived from the following virus strains: A/California/04/2009 (H1N1), A/Vietnam/1203/2004 (H5N1), A/California/135/2024 (H5N1) and A/Guangdong/17SF003/2016 (H7N9). Recombinant NA proteins (as soluble tetramers) used in ELISA were derived from the following isolates: A/California/04/2009 (H1N1), A/Vietnam/1203/2004 (H5N1), A/bald eagle/FL/W22-134-OP/2022 (H5N1) (B1.1 genotype,) A/California/135/2024 (H5N1) (B3.13 genotype(16)), A/Louisiana/12/2024 (H5N1) (D1.1 genotype), A/great black-backed gull/NS/FAV-0405-1/2023 (H5N5) (A6 genotype), A/Washington/2148/2025 (H5N5) (A6 genotype), A/Anhui/1/2013 (H7N9). The ectodomains of the HA and NA proteins were cloned into a baculovirus shuttle vector. The HA expression vector included a C-terminal trimerization domain and a hexahistidine tag whereas the NA vector contained an N-terminal tetramerization domain and hexahistidine tag. The baculoviruses were propagated in *Spodoptera frugiperda* (Sf9) cells, and protein expression was carried out in High Five cells (*Trichoplusia ni*). Briefly, High Five cells were infected with recombinant baculoviruses at a multiplicity of infection (MOI) of 10. Cell culture supernatants were then harvested by low-speed centrifugation 72 h post infection and were purified using Ni2+-nitrilotriacetic acid (Ni-NTA) chromatography. Protein purity and identity were tested by sodium dodecyl-sulfate polyacrylamide gel electrophoresis (SDS-PAGE) and Coomassie staining. Final protein concentrations were determined with Bradford reagent.

### Enzyme-linked immunosorbent assay (ELISA)

ELISA was performed for binding antibody response to different HA and NA antigens. In brief 96-well microtiter plates (Immulon-4 HBX; Thermo Fisher) were coated with 50 µL/well of the corresponding HA or NA antigens diluted at a concentration of 2 μg/ml and incubated at 4°C overnight. Plates were then washed the next day three times with phosphate-buffered saline (PBS) containing 0.1% Tween 20 (PBS-T). Blocking solution containing PBS-T, 3% goat serum and 0.5% milk powder was added to the plates (220 µL/well) and incubated for 1 h at room temperature (RT). Blocking solution was removed, samples were serially diluted 2-fold and added to the plates at a starting dilution of 1:100 in blocking solution (100 µL/well). Plates were incubated for 2 h at RT, then washed three times with PBS-T. The secondary anti-Human IgG (Fb specific)-Peroxidase antibody produced in goat (Sigma-Aldrich) was added at a volume of 50 µL/well, incubating for 1 h at RT. Plates were washed four times with PBS-T and developed with SigmaFast *o*-phenylenediamine dihydrochloride (OPD; Sigma) for 10 min at RT, then the reaction was stopped with 3M hydrochloric acid (Thermo Fisher Scientific). Using a Synergy H1 microplate reader (BioTek), the plates were read at an optical density (OD) of 490 nm. Background level was calculated as the average plus three times the standard deviation of blank wells in which no sample was added. Antibody levels expressed as the area under the curve (AUC) were calculated using Prism 10 (GraphPad). An anti-HA antibody (CR9114)(59) and an anti-H5 antibody (17E3)(60) were used as positive controls, while an anti–severe acute respiratory syndrome coronavirus 2 (SARS-CoV-2)(61) spike antibody was used as a negative control.

### Treatment of serum samples with receptor-destroying enzyme (RDE)

One volume of human sera was treated with three volumes of RDE from *Vibrio cholerae* (Denka Seiken, Tokyo, Japan) to remove non-specific inhibitors of hemagglutination and incubated at 37 °C water bath for 16-18 h. The following day, to the RDE-treated samples, three volumes of a 2.5% sodium citrate solution were added. After incubation at 56 °C for 1 h, three volumes of PBS were added to each sample for a final dilution of 1:10.

### Hemagglutinin inhibition assay (HI)

Sera were assessed for antibody to various influenza virus strains to determine antibody reaction to HA by HI assay using standard methods. A hemagglutination assay was initially performed to determine the hemagglutination titer units (HAU) of the viruses. RDE treated sera were serially diluted 1:2 in PBS in V-bottom 96-well plates. Twenty-five µL of serum dilutions were incubated with 25 µL of viruses diluted to 8 HAU at RT for 30 minutes (min) to allow HA-specific antibodies to bind to the virus. Then 50 µL of a 0.5% suspension of turkey red blood cells (Lampire) that was washed once with PBS were added to each well, and the plates were incubated at 4 °C until the red blood cells in PBS control samples settled to the bottom of the wells. HI titers were defined as the reciprocal of the highest dilution of serum that inhibited hemagglutination of red blood cells. Positive controls included the anti-H1-specific antibody 7B2(62) and the anti-H5-specific antibody 17E3(60), while an anti–SARS-CoV-2 spike(61) antibody was used as a negative control.

### Microneutralization (MNT) assay

The microneutralization assay was performed as described before(29). Briefly, MDCK cells were seeded in 96-well cell-culture treated plates (Corning) at density of 2.0 × 10^5^ cells/mL (100 µL/well) and incubated at 37 °C with 5% CO_2_ O/N. The following day, human sera treated with RDE was initially diluted 1:10 and serially diluted 2-fold across the plates in infection media consisting of 1x minimum essential media (MEM) (Gibco), 100 U/mL penicillin and 100 µg/mL streptomycin (Gibco), 10 mM 4-(2-hydroxyethyl)-1-piperazineethanesulfonic acid (HEPES) (Gibco), 2 mM L-glutamine (Gibco), 3.2% NaHCO_3_ (Sigma-Aldrich), and 1.2% bovine serum albumin (BSA) (MP Biomedicals) supplemented with 1 µg/mL N-tosyl-L-phenylalanine chloromethyl ketone (TPCK) -treated trypsin (Sigma-Aldrich). Next, 60 µL of 100 × 50% tissue culture infectious dose (TCID50) of virus prepared in infection medium and 60 µL of serially diluted sera were incubated on a shaker at RT for 1 h to allow antibodies to bind to virions. Before the end of the incubation, MDCK cells were washed with 220 µL of PBS and incubated with 100 µL of the incubated serum-virus mixture at 37 °C with 5% CO_2_ for 1 h to allow for attachment of virions to the cells. Afterwards, the virus inoculum was carefully aspirated, MDCK cells were washed with PBS, and 100 µL of the serially diluted sera containing TPCK treated trypsin were added to the cells and incubated at 37°C with 5% CO_2_ for 48h. This means the virus was allowed to replicate over several replication cycles with serum present at during the initial infection as well as the incubation period. As readout, the presence of virus was assessed by hemagglutination assay. In brief, 50 µL of cell supernatant was added to 96-well V bottom plates (Nunc) and serially diluted 1:2. Then, 50 µL of 0.5% v/v turkey red blood cells (RBCs, Lampire Biological Laboratories) in PBS were added to each well, and plates. Data are displayed as the endpoint titer and this value represents the lowest dilution at which no hemagglutination could be detected. An anti-HA antibody CR9114(59) was used as a positive control, while an anti–SARS-CoV-2 spike antibody(59) was used as a negative control.

### Neuraminidase inhibition (NI) assay

To measure NI activity, an enzyme linked lectin assay (ELLA) was performed to evaluate NA-inhibiting antibody titers in serum samples against H15N1_A/California/04/2009,_ H15N1_A/Louisiana/2024,_ H15N1_A/California/135/2024,_ H15N1_A/British Columbia/PHL-2032/2024_, A/bald eagle/FL/W22-134-OP/2022 (H5N1) A/Vietnam/1203/2004 (H5N1) and H15N5 _A/great black-backed gull/NS/FAV-0405-1/2023_ and H15N5 _A/Washington/2148/2025_ influenza viruses, following an established protocol(63). First, a NA assay was performed to determine the optimal virus concentration to be used in the NI assay. 96-well plates (Immulon-4 HBX, Thermo Scientific) were coated with 100 µL/well of fetuin (Sigma) at a concentration of 25 µg/mL in 1X PBS and stored at 4°C O/N. The next day, fetuin-coated plates were washed three times with PBS-T and blocked with PBS-T supplemented with 1% bovine serum albumin (BSA, MP Biomedicals). On a separate 96-well plate, virus was serially diluted 1:2 in PBS starting at a 1:10 initial dilution. 60 μL of PBS was added to all wells and the plates incubated for 1.5 h on a shaker at room temperature. After blocking, the plates were washed three times with PBS-T, 100 μL of serially diluted virus was transferred well by well to the fetuin coated plates, and the plates were incubated for O/N at 37°C. Afterwards, plates were washed six times with PBS-T, and 100 µL per well of peroxidase labeled peanut agglutinin from *Arachis hypogaea* (Millipore Sigma) at 5 µg/mL in PBS-T supplemented with 1% BSA was added to the plates. Plates were incubated at RT for 2 h before washing three times with PBS-T. To develop the plates, 100 μL of OPD substrate was added to each well. After a 10 min incubation, the reaction was stopped by adding 50 μL of 3 M HCl to each well. The OD_490_ was measured on a Synergy 4 plate reader (BioTek). The half maximal effective concentration (EC_50_) was determined using GraphPad Prism.

To perform NI assays, the microtiter plates were coated and blocked as described above. During the blocking of the fetuin-coated plates, the heat-treated sera (56 °C for 1 h) were diluted to a starting dilution of 1:10 and serially diluted 1:2 in PBS in a separate 96-well plate. 60 μL of virus diluted to 2 x half-maximal inhibitory concentration (IC_50_) was added to wells of the serially diluted sera and incubated for 1.5 h on a shaker. The fetuin-coated plates were washed three times with PBS-T, and the virus/sera mixture was added to the plates and incubated O/N at 37°C. The remainder of the assay was performed as described above. No serum (virus only) and background controls (PBS-T + 1% BSA only) were also included to measure the NI. OD490 was measured on a Synergy H1 microplate reader, and the half-maximal inhibitory concentration (IC_50_) was calculated as: 1 – (OD_measured_-OD_background_)/(OD_no serum control_ - OD _background_) in Microsoft Excel and GraphPad Prism 10. As a positive control anti-NA antibody, 1G01(63) was used, and as a negative control anti-SARS-CoV-2 spike antibody(59).

### Phylogenetic tree and model building

All HA and NA amino acid sequences categorized as H5N1, clade 2.3.4.4b, genotype D1.1 (GenoFlu genotyping) were downloaded on 14.0.2026 from Global Initiative on Sharing All Influenza Data (GISAID)(64). From all sets, all sequences containing “X” and exactly matching duplicates were removed. This resulted in 1772 of the available 3232 HA and 1864 of the available 3232 NA nucleotide sequences. As a rooting reference, the HA and NA nucleotide sequences from A/California/135/2024 (H5N1, clade 2.3.4.4b, genotype B3.13; EPI_ISL_19463618) were added to the sequence sets before creating the trees. Trees were built using the NGPhylogeny server (65) “A la carte” workflow, with MAFFT as alignment method(66), TrimAl for alignment curation(67), and FastTree for phylogenic tree inference(68–70). The trees were visualized using the iTOL server (71). The trees were rerooted to A/California/135/2024 sequences, and this branch was then removed from the tree for better visibility. Isolates in which the NA can be glycosylated at amino acid position 270 (N1 numbering) were marked with a lime green circle, and the ones in which this position is not a glycosylation site were marked with a purple square, based on the amino acid sequences. The position of the A/British Columbia/PHL-2032/2024 samples were marked with an orange star, including the A/British Columbia/PHL-2032-recombinant/2025 (EPI_ISL_19873158) sample. The closest sequence to the HA samples in the trees were A/chicken/Oregon/W250070001-2/2025 (EPI ISL 20267529, HA) and A/chicken/USA/000540-001/2025 (EPI ISL 19737061 HA). The closest NA sequence on the tree is A/great horned owl/USA/006101-001/2025 (EPI ISL 19832464 NA), a sample that is not glycosylated at position 270. The A/British Columbia/PHL-2032/2024 samples are nested among a branch with non-glycosylated samples, but the branch is poorly supported (bootstrap: 0.422), making the origin of the glycosylation status of A/British Columbia/PHL-2032/2024 ambigous. In addition, the samples from the Pacific Northwest (British Columbia, Canada; Oregon and Washington, USA) from October-December 2024 migratory season were marked with blue triangles outside the tree.

### Statistical analysis

All statistical analyses, AUC calculation, and IC_50_ were conducted with Graphpad Prism version 10. Differences between antibody titers between three groups were analyzed with a Kruskal–Wallis with Dunn’s multiple comparisons test. Correlations between antibody titers were analyzed using Spearman’s rank test. Significance was considered with p-values equal or less than 0.05 (*), ≥0.01 (**), ≥0.001 (***), ≥0.0001 (****). Smooth curves and confidence intervals calculated using the GLMGam module of statsmodels (v0.14.4) in Python (v3.12.2) with cubic BSplines using 6 degrees of freedom. Plots for these curves were generated using seaborn (v0.13.2).

## Data Availability

All data produced in the present study are available upon reasonable request to the authors.

## Acknowledgments

We thank all the study participants for their generous support of our research programs. Special thanks go to the teams of the APOLLO study and the Personalized Virology Initiative for sharing biospecimen and metadata from study participants. This work was funded through the NIAID Centers for Excellence in Influenza Research and Response (75N93021C00014 as well as option APOLLO Option 12A) and through philanthropic support from Tito’s Vodka. Work at the Medical University of Vienna was funded by Institutional Funds. We gratefully acknowledge all data contributors, i.e., the Authors and their Originating laboratories responsible for obtaining the specimens, and their Submitting laboratories for generating the genetic sequence and metadata and sharing via the GISAID Initiative, on which this research is based.

## Conflict of interest statement

The Icahn School of Medicine at Mount Sinai has filed patent applications relating to SARS-CoV-2 serological assays, NDV-based SARS-CoV-2 vaccines influenza virus vaccines and influenza virus therapeutics which list FK as co-inventor and FK has received royalty payments from some of these patents. Mount Sinai has spun out a company, Castlevax, to develop SARS-CoV-2 vaccines. VS is listed on the patent for the SARS-CoV-2 serological assay. FK is co-founder and scientific advisory board member of Castlevax. FK has consulted for Merck, GSK, Sanofi, Gritstone, Curevac, Seqirus and Pfizer and is currently consulting for 3rd Rock Ventures and Avimex. The Krammer laboratory is also collaborating with Dynavax on influenza vaccine development.

---

**Supplementary Fig. 1.**
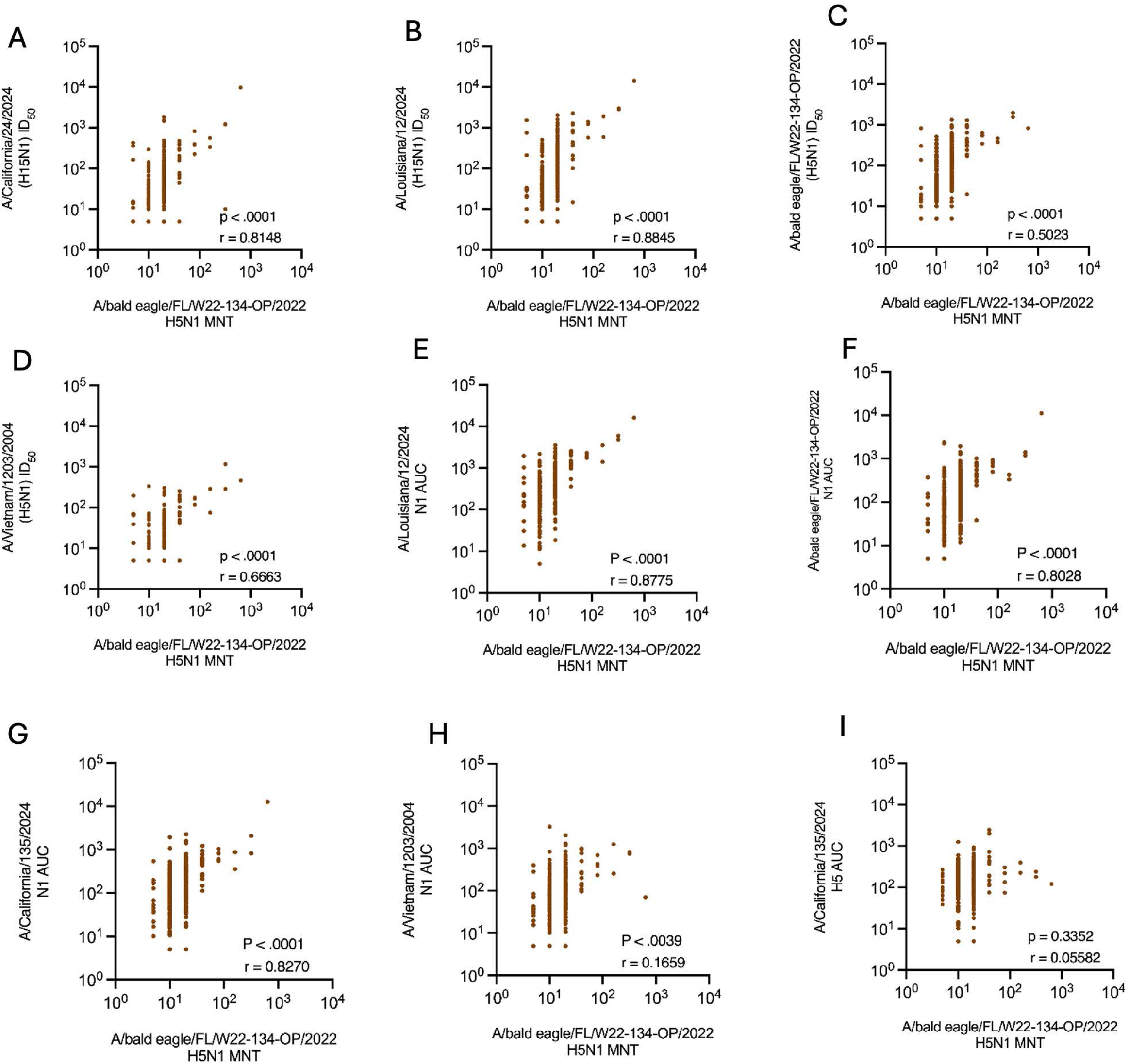
Spearmen correlation of H5N1 microneutralization and antigen specific binding and NI antibodies. (**A-D**) Spearmen correlation analysis was performed H5N1 microneutralization titers and H5N1 viruses NI antibodies (**E-H**) Spearmen correlation analysis was performed H5N1 microneutralization titers and N1 NA binding antibodies of H5N1 viruses. (I) Spearmen correlation analysis were performed H5N1 microneutralization titers and HA binding antibodies of H5N1 viruses. Correlation coefficient (r) and significance value (p) are indicated above the x axis.

**Supplementary Fig. 2.**
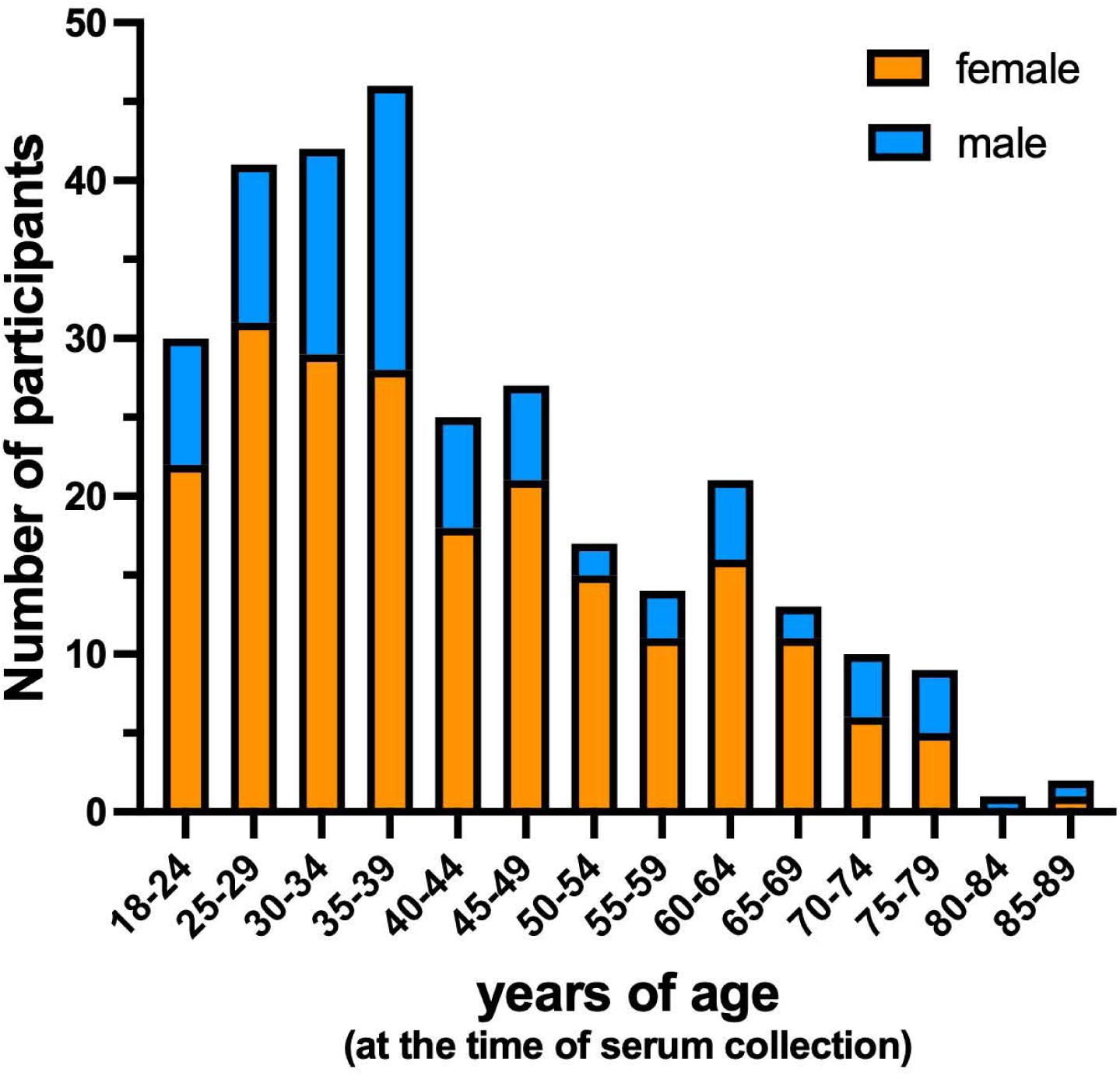
Age and sex distribution of individuals from whom sera were analyzed.

**Table S1.**
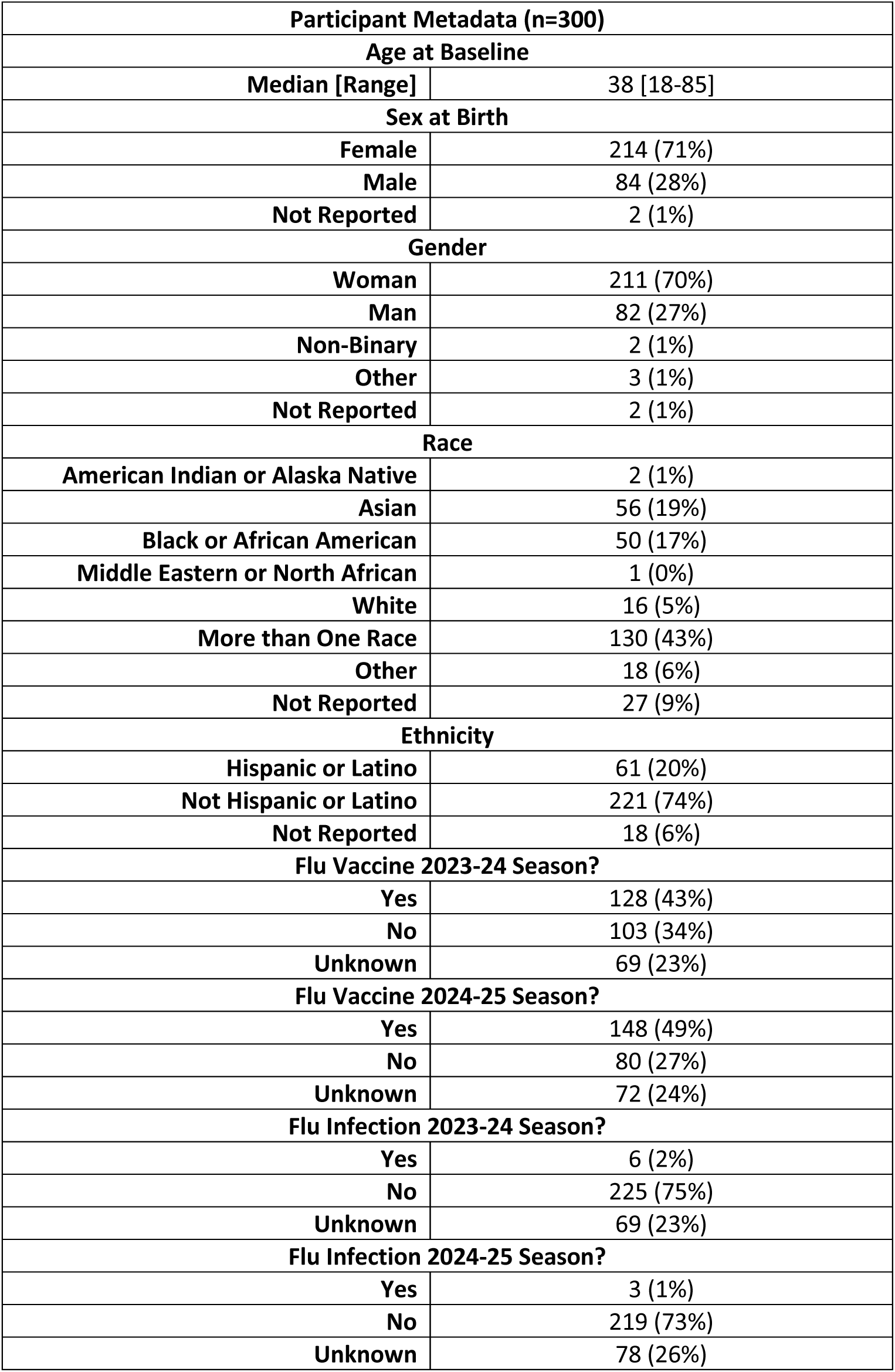
Detailed description of samples used (collected between February 2024 and April 2025)

**Table S2:** Percent amino acid identity of HA sequences of A(H5N1) and seasonal A(H1N1)

|  | **A/Cal/04/09**<br>**H1** | **A/VN/1203/04**<br>**H5**<br>**clade 1** | **A/Cal/135/24**<br>**H5 clade**<br>**2.3.4.4b** | **A/Guna/17SF003/16**<br>**H7** |
| --- | --- | --- | --- | --- |
| **A/Cal/04/09 H1** | 100 | 64.0 | 63.7 | 40.9 |
| **A/VN/1203/04 H5**<br>**clade 1** | 64.0 | 100 | 91.3 | 63.1 |
| **A/Cal/135/24 H5**<br>**2.3.4.4b** | 63.7 | 91.3 | 100 | 62.5 |
| **A/Guna/17SF003/16**<br>**H7** | 40.9 | 63.1 | 62.5 | 100 |

**Table S3:** Percent amino acid identity of NA sequences of A(H5N1) and seasonal A(H1N1)

|  | **A/Cal/**<br>**04/09**<br>**N1** | **A/VN/120**<br>**3/04 N1**<br>**clade 1** | **A/BE/FL/2**<br>**2 N1 clade**<br>**2.3.4.4b**<br>**(B1.1)** | **A/Cal/135/**<br>**24 N1**<br>**clade**<br>**2.3.4.4b**<br>**(B3.13)** | **A/LA/12/2**<br>**4 N1 clade**<br>**2.3.4.4b**<br>**(D1.1)** | **A/Gull/**<br>**NS/23**<br>**N5**<br>**(A6)** | **A/Wash**<br>**/2148/25**<br>**N5**<br>**(A6)** | **A/AH/1/13**<br>**N9** |
| --- | --- | --- | --- | --- | --- | --- | --- | --- |
| **A/Cal/04**<br>**/09 N1** | 100.0 | 72.3 | 78.3 | 78.3 | 83.3 | 59.1 | 59.1 | 46.0 |
| **A/VM/12**<br>**03/04 N1** | 72.3 | 100.0 | 87.5 | 87.5 | 83.0 | 61.0 | 61.0 | 47.2 |
| **A/BE/FL**<br>**/22 N1** | 78.3 | 87.5 | 100.0 | 99.0 | 88.3 | 63.0 | 63.0 | 47.2 |
| **A/Cal/13**<br>**5/24 N1** | 78.3 | 87.5 | 99.0 | 100.0 | 88.3 | 63.0 | 63.0 | 46.1 |
| **A/LA/12/**<br>**24 N1** | 83.3 | 83.0 | 88.3 | 88.3 | 100.0 | 62.5 | 62.5 | 46.2 |
| **A/Gull/N**<br>**S/23 N5** | 59.1 | 61.0 | 63.0 | 63.0 | 62.0 | 100.0 | 99.0 | 41.0 |
| **A/Wash/**<br>**2148/25**<br>**N5** | 59.1 | 61.0 | 63.0 | 63.0 | 62.5 | 99.0 | 100 | 41.0 |
| **A/AH/1/**<br>**13 N9** | 46.0 | 47.2 | 47.2 | 46.1 | 46.2 | 41.0 | 41.0 | 100 |

## Notes

### Author Declarations

Sera were collected between February 2024 and April 2025 from adult study participants who live or work in New York City as part of the observational study APOLLO (Antibody panels of longitudinal levels of coronavirus immunity) carried out at the ISMMS (Icahn School of Medicine at Mount Sinai). The study is approved by the Mount Sinai Hospital Institutional Review Board (STUDY-23-00873) and recorded as study protocol DMID 24-0011 as part of the Center for Research on Influenza Pathogenesis (CRIPT). All participants signed written consent forms prior to sample and data collection.

## References

1. De BK, Brownlee GG, Kendal AP, Shaw MW. 1988. Complete sequence of a cDNA clone of the hemagglutinin gene of influenza A/Chicken/Scotland/59 (H5N1) virus: comparison with contemporary North American and European strains. Nucleic Acids Res 16:4181–2.

2. Claas EC, Osterhaus AD, van Beek R, De Jong JC, Rimmelzwaan GF, Senne DA, Krauss S, Shortridge KF, Webster RG. 1998. Human influenza A H5N1 virus related to a highly pathogenic avian influenza virus. Lancet 351:472–7.

3. Yuen KY, Chan PK, Peiris M, Tsang DN, Que TL, Shortridge KF, Cheung PT, To WK, Ho ET, Sung R, Cheng AF. 1998. Clinical features and rapid viral diagnosis of human disease associated with avian influenza A H5N1 virus. Lancet 351:467–71.

4. Xu X, Subbarao, Cox NJ, Guo Y. 1999. Genetic characterization of the pathogenic influenza A/Goose/Guangdong/1/96 (H5N1) virus: similarity of its hemagglutinin gene to those of H5N1 viruses from the 1997 outbreaks in Hong Kong. Virology 261:15–9.

5. Krammer F, Hermann E, Rasmussen AL. 2025. Highly pathogenic avian influenza H5N1: history, current situation, and outlook. J Virol 99:e0220924.

6. Damodaran L, Jaeger AS, Moncla LH. 2025. Ecology and spread of the North American H5N1 epizootic. Nature.

7. Lair S, Quesnel L, Signore AV, Delnatte P, Embury-Hyatt C, Nadeau MS, Lung O, Ferrell ST, Michaud R, Berhane Y. 2024. Outbreak of Highly Pathogenic Avian Influenza A(H5N1) Virus in Seals, St. Lawrence Estuary, Quebec, Canada. Emerg Infect Dis 30:1133–1143.

8. Puryear W, Sawatzki K, Hill N, Foss A, Stone JJ, Doughty L, Walk D, Gilbert K, Murray M, Cox E, Patel P, Mertz Z, Ellis S, Taylor J, Fauquier D, Smith A, DiGiovanni RA, van de Guchte A, Gonzalez-Reiche AS, Khalil Z, van Bakel H, Torchetti MK, Lantz K, Lenoch JB, Runstadler J. 2023. Highly Pathogenic Avian Influenza A(H5N1) Virus Outbreak in New England Seals, United States. Emerg Infect Dis 29:786–791.

9. Uhart MM, Vanstreels RET, Nelson MI, Olivera V, Campagna J, Zavattieri V, Lemey P, Campagna C, Falabella V, Rimondi A. 2024. Epidemiological data of an influenza A/H5N1 outbreak in elephant seals in Argentina indicates mammal-to-mammal transmission. Nat Commun 15:9516.

10. Peacock TP, Moncla L, Dudas G, VanInsberghe D, Sukhova K, Lloyd-Smith JO, Worobey M, Lowen AC, Nelson MI. 2025. The global H5N1 influenza panzootic in mammals. Nature 637:304–313.

11. Caserta LC, Frye EA, Butt SL, Laverack M, Nooruzzaman M, Covaleda LM, Thompson AC, Koscielny MP, Cronk B, Johnson A, Kleinhenz K, Edwards EE, Gomez G, Hitchener G, Martins M, Kapczynski DR, Suarez DL, Alexander Morris ER, Hensley T, Beeby JS, Lejeune M, Swinford AK, Elvinger F, Dimitrov KM, Diel DG. 2024. Spillover of highly pathogenic avian influenza H5N1 virus to dairy cattle. Nature 634:669–676.

12. Halwe NJ, Cool K, Breithaupt A, Schön J, Trujillo JD, Nooruzzaman M, Kwon T, Ahrens AK, Britzke T, McDowell CD, Piesche R, Singh G, Pinho Dos Reis V, Kafle S, Pohlmann A, Gaudreault NN, Corleis B, Ferreyra FM, Carossino M, Balasuriya UBR, Hensley L, Morozov I, Covaleda LM, Diel DG, Ulrich L, Hoffmann D, Beer M, Richt JA. 2024. H5N1 clade 2.3.4.4b dynamics in experimentally infected calves and cows. Nature.

13. Rolfes MA, Kniss K, Kirby MK, Garg S, Reinhart K, Davis CT, Murray EL, Wadford DA, Harriman K, Zhu S, Liu CY, Morales C, Lopez D, Esbenshade L, Trivedi KK, Tobolowsky FA, Dang T, Unutzer A, Krager S, Baum K, Curren EJ, Harrist A, Hand J, Sokol T, Salinas A, Bunch A, Haydel D, Fisher N, Margrey SF, Billing L, Turabelidze G, Goswitz J, Kavlak L, Gonzales E, Tillman W, Rollo S, Hamid CA, Aragon D, Owen M, Kohnen A, Hoehne A, Weigel A, Hennenfent A, Sepcic V, Southern T, Beard C, Scholz R, Present S, Sutton M, Morse J, et al. 2025. Human infections with highly pathogenic avian influenza A(H5N1) viruses in the United States from March 2024 to May 2025. Nat Med 31:3889–3898.

14. Kandeil A, Patton C, Jones JC, Jeevan T, Harrington WN, Trifkovic S, Seiler JP, Fabrizio T, Woodard K, Turner JC, Crumpton JC, Miller L, Rubrum A, DeBeauchamp J, Russell CJ, Govorkova EA, Vogel P, Kim-Torchetti M, Berhane Y, Stallknecht D, Poulson R, Kercher L, Webby RJ. 2023. Rapid evolution of A(H5N1) influenza viruses after intercontinental spread to North America. Nat Commun 14:3082.

15. Signore AV, Giacinti J, Jones MEB, Erdelyan CNG, McLaughlin A, Alkie TN, Cox S, Lair S, Jardine CM, Stevens B, Bravo-Araya M, Pople N, Pybus MJ, Hisanaga T, Xu W, Koziuk J, Lung O, Kruczkiewicz P, Fisher M, Wight J, Rahman I, Hargan KE, Lang AS, Hochman O, Ojkic D, Yason C, Bourque L, Bollinger TK, Provencher JF, Ogilvie S, Clark A, MacPhee R, Eaglesome H, Gilbert S, Saboraki K, Davis R, Jerao A, Ginn M, Soos C, Berhane Y, Program BCWAS. 2025. Spatiotemporal reconstruction of the North American A(H5N1) outbreak reveals successive lineage replacements by descendant reassortants. Sci Adv 11:eadu4909.

16. Youk S, Torchetti MK, Lantz K, Lenoch JB, Killian ML, Leyson C, Bevins SN, Dilione K, Ip HS, Stallknecht DE, Poulson RL, Suarez DL, Swayne DE, Pantin-Jackwood MJ. 2023. H5N1 highly pathogenic avian influenza clade 2.3.4.4b in wild and domestic birds: Introductions into the United States and reassortments, December 2021-April 2022. Virology 587:109860.

17. Crespo-Bellido A, Trovão NS, Maksiaev A, Baele G, Dellicour S, Nelson MI. 2025. Emergence of D1.1 reassortant H5N1 avian influenza viruses in North America. bioRxiv:2025.12.19.695329.

18. Kendal AP, Joseph JM, Kobayashi G, Nelson D, Reyes CR, Ross MR, Sarandria JL, White R, Woodall DF, Noble GR, Dowdle WR. 1979. Laboratory-based surveillance of influenza virus in the United States during the winter of 1977-1978. I. Periods of prevalence of H1N1 and H3N2 influenza A strains, their relative rates of isolation in different age groups, and detection of antigenic variants. Am J Epidemiol 110:449–61.

19. Pyankova OG, Susloparov IM, Moiseeva AA, Kolosova NP, Onkhonova GS, Danilenko AV, Vakalova EV, Shendo GL, Nekeshina NN, Noskova LN, Demina JV, Frolova NV, Gavrilova EV, Maksyutov RA, Ryzhikov AB. 2021. Isolation of clade 2.3.4.4b A(H5N8), a highly pathogenic avian influenza virus, from a worker during an outbreak on a poultry farm, Russia, December 2020. Euro Surveill 26.

20. Jassem AN, Roberts A, Tyson J, Zlosnik JEA, Russell SL, Caleta JM, Eckbo EJ, Gao R, Chestley T, Grant J, Uyeki TM, Prystajecky NA, Himsworth CG, MacBain E, Ranadheera C, Li L, Hoang LMN, Bastien N, Goldfarb DM. 2025. Critical Illness in an Adolescent with Influenza A(H5N1) Virus Infection. N Engl J Med 392:927–929.

21. Taylor L. 2025. Man in first ever human case of H5N5 avian flu dies. BMJ 391:r2480.

22. Krammer F, Smith GJD, Fouchier RAM, Peiris M, Kedzierska K, Doherty PC, Palese P, Shaw ML, Treanor J, Webster RG, García-Sastre A. 2018. Influenza. Nat Rev Dis Primers 4:3.

23. Sidney J, Kim AR, de Vries RD, Peters B, Meade PS, Krammer F, Grifoni A, Sette A. 2025. Targets of influenza human T-cell response are mostly conserved in H5N1. mBio 16:e0347924.

24. Power MA, Rijnink WF, Soochit W, Gommers L, van der Linden A, Chandler F, Volker F, Bestebroer TM, Verstrepen BE, Grifoni A, Tan NH, Bogers S, van Nierop GP, Sette A, Koopmans MPG, GeurtsvanKessel CH, Sikkema RS, Richard M, de Vries RD. 2025. Seasonal Influenza Exposure Elicits Functional Antibody and T-cell Responses to A(H5) Influenza Viruses in Humans. medRxiv:2025.07.31.25331995.

25. Nachbagauer R, Choi A, Hirsh A, Margine I, Iida S, Barrera A, Ferres M, Albrecht RA, García-Sastre A, Bouvier NM, Ito K, Medina RA, Palese P, Krammer F. 2017. Defining the antibody cross-reactome directed against the influenza virus surface glycoproteins. Nat Immunol.

26. Erdelyan CNG, Kandeil A, Signore AV, Jones MEB, Vogel P, Andreev K, Bøe CA, Gjerset B, Alkie TN, Yason C, Hisanaga T, Sullivan D, Lung O, Bourque L, Ayilara I, Pama L, Jeevan T, Franks J, Jones JC, Seiler JP, Miller L, Mubareka S, Webby RJ, Berhane Y. 2024. Multiple transatlantic incursions of highly pathogenic avian influenza clade 2.3.4.4b A(H5N5) virus into North America and spillover to mammals. Cell Rep 43:114479.

27. Krammer F, Weir JP, Engelhardt O, Katz JM, Cox RJ. 2019. Meeting report and review: Immunological assays and correlates of protection for next-generation influenza vaccines. Influenza Other Respir Viruses.

28. Hobson D, Curry RL, Beare AS, Ward-Gardner A. 1972. The role of serum haemagglutination-inhibiting antibody in protection against challenge infection with influenza A2 and B viruses. J Hyg (Lond) 70:767–77.

29. Cuevas F, Kawabata H, Krammer F, Carreño JM. 2022. An In Vitro Microneutralization Assay for Influenza Virus Serology. Curr Protoc 2:e465.

30. Couch RB, Atmar RL, Franco LM, Quarles JM, Wells J, Arden N, Niño D, Belmont JW. 2013. Antibody correlates and predictors of immunity to naturally occurring influenza in humans and the importance of antibody to the neuraminidase. J Infect Dis 207:974–81.

31. Monto AS, Petrie JG, Cross RT, Johnson E, Liu M, Zhong W, Levine M, Katz JM, Ohmit SE. 2015. Antibody to Influenza Virus Neuraminidase: An Independent Correlate of Protection. J Infect Dis 212:1191–9.

32. Krammer F, Fouchier RAM, Eichelberger MC, Webby RJ, Shaw-Saliba K, Wan H, Wilson PC, Compans RW, Skountzou I, Monto AS. 2018. NAction! How Can Neuraminidase-Based Immunity Contribute to Better Influenza Virus Vaccines? MBio 9.

33. Monto AS, Kendal AP. 1973. Effect of neuraminidase antibody on Hong Kong influenza. Lancet 1:623–5.

34. Zhang L, Behrens GMN, Kempf A, Nehlmeier I, Gärtner S, Moldenhauer AS, Graichen L, Happle C, Winkler M, Dopfer-Jablonka A, Pöhlmann S, Hoffmann M. 2025. Neutralizing activity against bovine H5N1 HPAIV (clade 2.3.4.4b) in human plasma after seasonal influenza vaccination. Emerg Microbes Infect 14:2528539.

35. Daniel K, Ullrich L, Ruchnewitz D, Meijers M, Halwe NJ, Wild U, Eberhardt J, Schön J, Stumpf R, Schlotz M, Wunsch M, Girao Lessa L, Abd El-Whab E-SM, Kuryshko M, Dietrich C, Pinger A, Schumacher A-L, Germer M, Rohde M, Kukat C, Gieselmann L, Gruell H, Hoffmann D, Beer M, Erren T, Lässig M, Kreer C, Klein F. 2025. Detection of pre-existing humoral immunity against influenza virus H5N1 clade 2.3.4.4b in unexposed individuals. bioRxiv:2025.01.22.634277.

36. Garretson TA, Liu J, Li SH, Scher G, Santos JJS, Hogan G, Vieira MC, Furey C, Atkinson RK, Ye N, Ort JT, Kim K, Hernandez KA, Eilola T, Schultz DC, Cherry S, Cobey S, Hensley SE. 2025. Immune history shapes human antibody responses to H5N1 influenza viruses. Nat Med 31:1454–1458.

37. Li ZN, Liu F, Jung YJ, Jefferson S, Holiday C, Gross FL, Tzeng WP, Carney P, Kates A, York IA, Safdar N, Zhou J, Medina MJ, Cioce V, Oshansky CM, Todd Davis C, Stevens J, Tumpey T, Levine MZ. 2025. Pre-existing cross-reactive immunity to highly pathogenic avian influenza 2.3.4.4b A(H5N1) virus in the United States. Nat Commun 16:10954.

38. Sanz-Muñoz I, Sánchez-Martínez J, Rodríguez-Crespo C, Concha-Santos CS, Hernández M, Rojo-Rello S, Domínguez-Gil M, Mostafa A, Martinez-Sobrido L, Eiros JM, Nogales A. 2025. Are we serologically prepared against an avian influenza pandemic and could seasonal flu vaccines help us? mBio 16:e0372124.

39. Daulagala P, Cheng SMS, Chin A, Luk LLH, Leung K, Wu JT, Poon LLM, Peiris M, Yen HL. 2024. Avian Influenza A(H5N1) Neuraminidase Inhibition Antibodies in Healthy Adults after Exposure to Influenza A(H1N1)pdm09. Emerg Infect Dis 30:168–171.

40. Ort JT, Leonard AS, Li SH, Atkinson RK, Mendoza LM, Vieira MC, Gang S, Cobey S, Hensley SE. 2025. Cross-reactive human antibody responses to H5N1 influenza virus neuraminidase are shaped by immune history. medRxiv.

41. Skowronski DM, Ranadheera C, Kaweski SE, Sabaiduc S, Separovic L, Makowski G, Ung J, Reyes RC, Henry B, Albert A, Clemens F, Kobasa D, Bastien N. 2026. Cross-reactive H5N1 neuraminidase antibodies by age and influenza A imprinting cohorts of the past century: population-based serosurvey, British Columbia, Canada. J Infect Dis.

42. Nachbagauer R, Choi A, Izikson R, Cox MM, Palese P, Krammer F. 2016. Age Dependence and Isotype Specificity of Influenza Virus Hemagglutinin Stalk-Reactive Antibodies in Humans. MBio 7.

43. Carnell GW, Ferrara F, Grehan K, Thompson CP, Temperton NJ. 2015. Pseudotype-based neutralization assays for influenza: a systematic analysis. Front Immunol 6:161.

44. Matsuda K, Huang J, Zhou T, Sheng Z, Kang BH, Ishida E, Griesman T, Stuccio S, Bolkhovitinov L, Wohlbold TJ, Chromikova V, Cagigi A, Leung K, Andrews S, Cheung CSF, Pullano AA, Plyler J, Soto C, Zhang B, Yang Y, Joyce MG, Tsybovsky Y, Wheatley A, Narpala SR, Guo Y, Darko S, Bailer RT, Poole A, Liang CJ, Smith J, Alexander J, Gurwith M, Migueles SA, Koup RA, Golding H, Khurana S, McDermott AB, Shapiro L, Krammer F, Kwong PD, Connors M. 2019. Prolonged evolution of the memory B cell response induced by a replicating adenovirus-influenza H5 vaccine. Sci Immunol 4.

45. Hermann E, Krammer F. 2025. Clade 2.3.4.4b H5N1 neuraminidase has a long stalk, which is in contrast to most highly pathogenic H5N1 viruses circulating between 2002 and 2020. mBio 16:e0398924.

46. Drake JM. 2025. Understanding avian influenza mortality. Science 389:1292–1294.

47. Wang TT, Parides MK, Palese P. 2012. Seroevidence for H5N1 influenza infections in humans: meta-analysis. Science 335:1463.

48. Le Sage V, Werner BD, Merrbach GA, Petnuch SE, O’Connell AK, Simmons HC, McCarthy KR, Reed DS, Moncla LH, Bhavsar D, Krammer F, Crossland NA, McElroy AK, Duprex WP, Lakdawala SS. 2025. Influenza A(H5N1) Immune Response among Ferrets with Influenza A(H1N1)pdm09 Immunity. Emerg Infect Dis 31:477–487.

49. Restori KH, Weaver V, Patel DR, Merrbach GA, Septer KM, Field CJ, Bernabe MJ, Kronthal EM, Minns A, Lindner SE, Lakdawala SS, Le Sage V, Sutton TC. 2025. Preexisting immunity to the 2009 pandemic H1N1 virus reduces susceptibility to H5N1 infection and disease in ferrets. Sci Transl Med 17:eadw4856.

50. Martínez-Sobrido L, García-Sastre A. 2010. Generation of recombinant influenza virus from plasmid DNA. J Vis Exp.

51. Beare AS, Schild GC, Craig JW. 1975. Trials in man with live recombinants made from A/PR/8/34 (H0 N1) and wild H3 N2 influenza viruses. Lancet 2:729–32.

52. Belser JA, Johnson A, Pulit-Penaloza JA, Pappas C, Pearce MB, Tzeng WP, Hossain MJ, Ridenour C, Wang L, Chen LM, Wentworth DE, Katz JM, Maines TR, Tumpey TM. 2017. Pathogenicity testing of influenza candidate vaccine viruses in the ferret model. Virology 511:135–141.

53. Campbell PJ, Danzy S, Kyriakis CS, Deymier MJ, Lowen AC, Steel J. 2014. The M segment of the 2009 pandemic influenza virus confers increased neuraminidase activity, filamentous morphology, and efficient contact transmissibility to A/Puerto Rico/8/1934-based reassortant viruses. J Virol 88:3802–14.

54. Rodriguez A, Pérez-González A, Hossain MJ, Chen LM, Rolling T, Pérez-Breña P, Donis R, Kochs G, Nieto A. 2009. Attenuated strains of influenza A viruses do not induce degradation of RNA polymerase II. J Virol 83:11166–74.

55. Matsuoka Y, Chen H, Cox N, Subbarao K, Beck J, Swayne D. 2003. Safety evaluation in chickens of candidate human vaccines against potential pandemic strains of influenza. Avian Dis 47:926–30.

56. Margine I, Palese P, Krammer F. 2013. Expression of functional recombinant hemagglutinin and neuraminidase proteins from the novel H7N9 influenza virus using the baculovirus expression system. J Vis Exp:e51112.

57. Krammer F, Margine I, Tan GS, Pica N, Krause JC, Palese P. 2012. A carboxy-terminal trimerization domain stabilizes conformational epitopes on the stalk domain of soluble recombinant hemagglutinin substrates. PLoS One 7:e43603.

58. Loganathan M, Francis B, Krammer F. 2024. Production of Influenza Virus Glycoproteins Using Insect Cells. Methods Mol Biol 2762:43–70.

59. Dreyfus C, Laursen NS, Kwaks T, Zuijdgeest D, Khayat R, Ekiert DC, Lee JH, Metlagel Z, Bujny MV, Jongeneelen M, van der Vlugt R, Lamrani M, Korse HJ, Geelen E, Sahin Ö, Sieuwerts M, Brakenhoff JP, Vogels R, Li OT, Poon LL, Peiris M, Koudstaal W, Ward AB, Wilson IA, Goudsmit J, Friesen RH. 2012. Highly conserved protective epitopes on influenza B viruses. Science 337:1343–8.

60. Alzua GP, León AN, Yellin T, Bhavsar D, Loganathan M, Bushfield K, Brouwer PJM, Rodriguez AJ, Jeevan T, Webby R, Marizzi C, Han J, Ward AB, Duty JA, Krammer F. 2025. Human monoclonal antibodies that target clade 2.3.4.4b H5N1 hemagglutinin. Nat Commun 17:135.

61. Tian X, Li C, Huang A, Xia S, Lu S, Shi Z, Lu L, Jiang S, Yang Z, Wu Y, Ying T. 2020. Potent binding of 2019 novel coronavirus spike protein by a SARS coronavirus-specific human monoclonal antibody. Emerg Microbes Infect 9:382–385.

62. Tan GS, Krammer F, Eggink D, Kongchanagul A, Moran TM, Palese P. 2012. A pan-H1 anti-hemagglutinin monoclonal antibody with potent broad-spectrum efficacy in vivo. J Virol 86:6179–88.

63. Stadlbauer D, McMahon M, Turner HL, Zhu X, Wan H, Carreño JM, O’Dell G, Strohmeier S, Khalil Z, Luksza M, van Bakel H, Simon V, Ellebedy AH, Wilson IA, Ward AB, Krammer F. 2022. Antibodies targeting the neuraminidase active site inhibit influenza H3N2 viruses with an S245N glycosylation site. Nat Commun 13:7864.

64. Khare S, Gurry C, Freitas L, Schultz MB, Bach G, Diallo A, Akite N, Ho J, Lee RT, Yeo W, Curation Team GC, Maurer-Stroh S. 2021. GISAID’s Role in Pandemic Response. China CDC Wkly 3:1049–1051.

65. Lemoine F, Correia D, Lefort V, Doppelt-Azeroual O, Mareuil F, Cohen-Boulakia S, Gascuel O. 2019. NGPhylogeny.fr: new generation phylogenetic services for non-specialists. Nucleic Acids Res 47:W260–W265.

66. Katoh K, Standley DM. 2013. MAFFT multiple sequence alignment software version 7: improvements in performance and usability. Mol Biol Evol 30:772–80.

67. Capella-Gutiérrez S, Silla-Martínez JM, Gabaldón T. 2009. trimAl: a tool for automated alignment trimming in large-scale phylogenetic analyses. Bioinformatics 25:1972–3.

68. Lemoine F, Domelevo Entfellner JB, Wilkinson E, Correia D, Dávila Felipe M, De Oliveira T, Gascuel O. 2018. Renewing Felsenstein’s phylogenetic bootstrap in the era of big data. Nature 556:452–456.

69. Price MN, Dehal PS, Arkin AP. 2010. FastTree 2--approximately maximum-likelihood trees for large alignments. PLoS One 5:e9490.

70. Price MN, Dehal PS, Arkin AP. 2009. FastTree: computing large minimum evolution trees with profiles instead of a distance matrix. Mol Biol Evol 26:1641–50.

71. Letunic I, Bork P. 2024. Interactive Tree of Life (iTOL) v6: recent updates to the phylogenetic tree display and annotation tool. Nucleic Acids Res 52:W78–W82.

